# Racialised Inequities in Career Progression among the Healthcare Workforce: A Qualitative Study

**DOI:** 10.64898/2026.06.17.26355685

**Authors:** Nathan M. Stanley, Dorothy Williams, Charlotte Woodhead, Nkasi Stoll, Amy Morgan, Cerisse Gunasinghe, Anna Ehsan, Juliana Onwumere, Anna-Theresa Jieman, Paula Meriez, Farah Ahmed, Stephani L. Hatch

## Abstract

**Aims:** To explore how workplace environments and racialised hierarchies shape racial inequities in career progression and how the Covid-19 pandemic response influenced these inequities.

**Design:** Longitudinal and cross-sectional qualitative study using semi-structured interviews.

**Methods:** Semi-structured interviews were conducted with 27 student nurses, healthcare assistants and qualified nurses and 24 senior leaders and management staff recruited in England between January 2019 and March 2021. Data were analysed using thematic analysis.

**Results:** Data from 51 healthcare professionals were included in the analysis. Guided by sociological theory on inequality diversions and racialised organisations, three main themes were identified: (1) ‘hidden pipelines to career progression’ highlights how racial positioning influences unequal career trajectories shaped by informal networks, organisational norms and perceptions of competence; (2) ‘impact of the response to the Covid-19 pandemic’ illustrates how the pandemic disrupted and reinforced racialised career barriers and (3) ‘psychological effects of racialised inequities on racially minoritised staff’ captures the emotional burden of navigating these inequities.

**Conclusion:** NHS staff perspectives on racialised inequities in career progression highlight the power informal networks have in staff accessing managers who control opportunities. While some staff found new opportunities during the Covid-19 response, others, particularly senior racially minoritised staff, felt redeployments and remote working further hindered their development.

**Impact:** Persistent racial inequities in NHS career progression must be addressed to improve staff retention and psychological wellbeing. Repeated exclusion and stalled progression, while advantaging white staff, negatively impacts racially minoritised staff, leading to fatigue, pessimism and reduced staff retention. Tackling these inequities requires confronting structural advantages for white staff, implementing mandatory anti-racism training for managers, carrying out robust evaluations of current organisational processes, embedding inclusive practices and offering targeted psychological support for racially minoritised staff.

**Patient or Public Contribution:** Input from national advisory and stakeholder opinion groups of healthcare professionals.

## 1 Introduction

### Inequalities in NHS Career Development and Progression

The National Health Service (NHS) is the UK’s biggest and most racially diverse employer, with 29% of staff from a racially minoritised background, groups actively marginalised due to their racial identities with ongoing underrepresentation at senior pay bands (Race Equality Action Group Queen Mary University of London, 2024; Rolewicz, Palmer and Lobont, 2022; NHS England, 2025; NHS, 2023) According to the 2024 Workforce Race Equality Standard (WRES) report, White NHS applicants were 1.6 times more likely to be appointed from shortlisting than racially minoritised staff across all posts (NHS England, 2025). In 2024, only 17% of NHS board members and less than 14% of staff at pay bands 8c and above (senior) were from a racially minoritised background (NHS England, 2025).

These racialised inequities are reflected in staff opinions: in the 2024 NHS staff survey, only 49% of racially minoritised staff believed that the NHS provides equal opportunities for career development, progression or promotion, mentorship, access to meetings and inclusion in senior spaces, compared to 59% of White staff (NHS England, 2024). Furthermore, qualitative studies have identified multiple organisational-level factors accounting for existing barriers to career development and promotion opportunities, including limited opportunities for skills development, training and mentorship (Isaac, 2020; Hammond et al., 2022; Williams et al., 2023). A recent review confirmed these are globally relevant findings and emphasised interpersonal barriers, such as these staff constantly having their authority undermined by White colleagues (Mostafa, Onwumere and Wood, 2025).

## 2 Background

Racialised inequities in career development and promotion are shaped by, organisational culture, norms and practices, which can create conditions for workplace bullying, harassment and discrimination (Jones, 2018; Rhead et al., 2021). UK-based qualitative findings suggest that staff lower in workforce hierarchies are more exposed to these adverse experiences, making them feel less valued (Walker et al., 2024; Woodhead et al., 2022). Racially minoritised staff often experience racial prejudice, stereotyping and exclusionary organisational norms. Staff responses to, and ways of coping with, these experiences led to workplace segregation and many racially minoritised staff changing teams or leaving the NHS entirely (Woodhead et al. 2022). Inevitably, experiences of bullying, harassment and discrimination in the NHS are associated with lower employee morale, productivity and poorer mental health and wellbeing (Rhead et al., 2021; Rhead et al., 2024).

Racialised inequities within the NHS are at least partially influenced by preference for specific racialised attributes and currency in having strong social networks within the NHS (Dhaliwal and McKay, 2017; Isaac, 2020). Black British-born mental healthcare staff shared their observation that NHS staff with English language proficiency, a British accent, British cultural norms and British education were more likely to progress in their NHS careers (Isaac (2020). Lacking these attributes appeared to lead to Black and other racially minoritised NHS healthcare staff being labelled by their colleagues and managers as unprofessional, incompetent and lacking motivation to pursue professional development or promotions (Dhaliwal and McKay, 2017; Isaac, 2020; Likupe et al., 2014).

## 3 The Study

### 3.1 Aims and Research Questions

This paper builds on prior findings about the influence of the NHS workplace context to explore, in greater depth and with stakeholder input, how the NHS workplace and racialised hierarchy affect career progression and subsequently, staff morale and psychological wellbeing. Notably, data collection coincided with the Covid-19 pandemic which significantly altered workplace pressures and career progression (Nicols et al., 2020; The King’s Fund, 2023). Therefore, this study also aims to explore how the pandemic response impacted racial and ethnic inequities in career development, including shifts in practices that may have reinforced or disrupted organisational norms within the NHS.

This study seeks to answer three questions: (1) How do health service organisational practices and norms affect racially minoritised staff’s ability to progress in their career? (2) What role do majority White staff and organisational Whiteness have in maintaining racial inequalities in the NHS? (3) In what ways, if any, did the response to the Covid-19 pandemic impact racialised inequities in career progression?

## 4 Methods

### 4.1 Design

This qualitative study used interview data from phase 1 (pre-Covid-19) and phase 2 (after the outbreak of Covid-19) of the Tackling Inequalities and Discrimination Experiences in Health Services (TIDES) study (tidesstudy.co.uk). The TIDES study aims to understand how discrimination experiences contribute to inequalities in health and health services. Full details of the methods and sample description were previously reported (Rhead et al., 2021; Woodhead et al., 2021).

### 4.2 Theoretical Framework

A key, underexplored area within this research is understanding the role of groups with privilege within the NHS workforce in maintaining inequities (e.g., Woodhead et al., 2022; Williams et al., 2023). Link and García (2021) identify ‘health inequality diversions’ in which the focus is often shifted away from “the health-inequality-generating actions of advantaged groups” (p. 346) to the characteristics of disadvantaged groups. This shift, labelled as a ‘diversion’, provides a deficit narrative about those who experience health inequalities. By diverting focus away from advantaged groups, they are protected from scrutiny and can continue health-inequality-generating actions (Link and García, 2021). Applied more broadly, such as to career progression, there is a need to acknowledge advantages White staff groups maintain, alongside disadvantages that racially minoritised staff face within organisations.

The current sub-study also draws on Ray’s Theory of Racialised Organisations, which posits that organisations are structures that support or challenge racialisation processes (Ray, 2019). Further, Ray (2019) argues that organisations enhance or diminish the agency of racial groups, legitimise unequal distribution of resources, racialise the decoupling of formal rules from organisational practice and hold Whiteness as a valuable credential and an organisational norm. Whiteness is defined as a culture centred on socially constructed power that is unequally distributed by skin colour (Chen, 2017; Hill, 1998; Trechter and Bucholtz, 2001).

### 4.3 Study Setting and Recruitment

The current sub-study analysis includes participants in the healthcare professionals (HCP) sample recruited in phase 1 and 2, as well as the senior leaders and management staff sample recruited in phase 2. The HCP sample was recruited as part of phase 1 of the TIDES study (Rhead et al., 2021) and were followed up in phase 2 after the onset of the pandemic. These participants initially completed a survey (n=931) and those who consented to be re-contacted were also purposively sampled for interviews. Of the 225 staff re-contacted, 48 (6 healthcare assistants, 21 student nurses, 10 entry-level nurses, and 11 mid-or senior-level nurses) participated in phase 1 interviews. Participants in the Senior Leaders and Managers sample (n=24) were recruited through a combination of snowballing and key informant sampling through the TIDES national advisory and stakeholder opinion groups. Interested participants were sent a participant information sheet with details on confidentiality and how to withdraw from the study. Participants received a £15 voucher to thank them their participation.

### 4.4 Inclusion and/or Exclusion Criteria

Inclusion criteria for the current sub-study analysis included: (1) student nurses, healthcare assistants and qualified nurses who were interviewed at both phase 1 and phase 2 (27 of the 48 interviewed at phase 1 were eligible) and (2) senior leaders and managers from phase 2 (n=24) who held senior-level management positions, senior-level clinicians with management responsibilities, or non-clinical senior-level staff in equality, diversity and inclusion-related roles from across 15 NHS Trusts in England.

### 4.5 Data collection

Data collection occurred between January 2019 and March 2021, starting before the pandemic and completed during England’s second national lockdown period. Phase 1 and phase 2 interviews were conducted by experienced qualitative researchers within the TIDES team and a TIDES collaborator, Challenge Consultancy (https://www.challcon.com/) also conducted interviews with the senior leaders and managers in phase 2. Data were transcribed verbatim, and any personal or organisationally identifying information was removed.

In phase 1, semi-structured interviews lasting 45-60 minutes were conducted either in-person or via telephone and were audio-recorded between January 2019 and February 2020. Phase 1 topic guides were developed through examining the literature and engaging with nurses and healthcare assistants at participating NHS Trusts. Topics included experiences of and witnessing workplace discrimination, bullying and harassment, reporting processes, training, support and retention.

For phase 2, semi-structured interviews lasting 45-60 minutes were conducted either online or via telephone and audio-recorded. Data were collected between October 2020 and March 2021, when Covid-19 infection, hospitalisation and death rates rapidly rose across the UK, putting unprecedented strain on health and social care services. This was a period of drastic change for NHS staff, and it impacted many different aspects of their lives at work and at home (Nichols et al., 2020). Topic guides were developed through a modified Delphi consensus process, with the input of national advisory and stakeholder opinion groups and refined in line with pandemic developments. Topics included changes to roles and responsibilities since the pandemic, access to personal protective equipment, vaccine perceptions, and experiences and witnessing of workplace discrimination, bullying and harassment.

### 4.6 Data Analysis

Transcripts from interviews with 27 HCP participants (interviewed at phase 1 and phase 2) and 24 senior leaders and managers were analysed. Thematic analysis (Braun and Clarke, 2006) was undertaken by racially and ethnically inclusive researchers within the TIDES team. Following familiarisation with the data, inductive, semantic and latent coding techniques were used to code transcripts by one researcher (NMS) to develop an initial analytical framework. Two other researchers (AM and NS) also descriptively hand coded, via MS Word, a sample of the transcripts, which allowed the analytical framework to be iteratively refined through repeated rounds of collaborative discussion (NMS, AM, NS, CW) and coding. The resulting themes were then presented to the wider group of co-authors, including NHS peer researchers with nursing backgrounds, to further refine the analytical framework. Data were imported into NVivo 11 (QSR International Pty Ltd. [2015] NVivo Version 11) and the analytical framework was applied to all transcripts. With no major difference in interpretation, any nuances were deliberated through discussion, and codes were grouped together to develop themes and sub-themes. Through an iterative process involving frequent cross-checking with transcript data, codes, themes and sub-themes were appraised and adjusted with links between themes examined (Figure 1).

**Figure 1.**
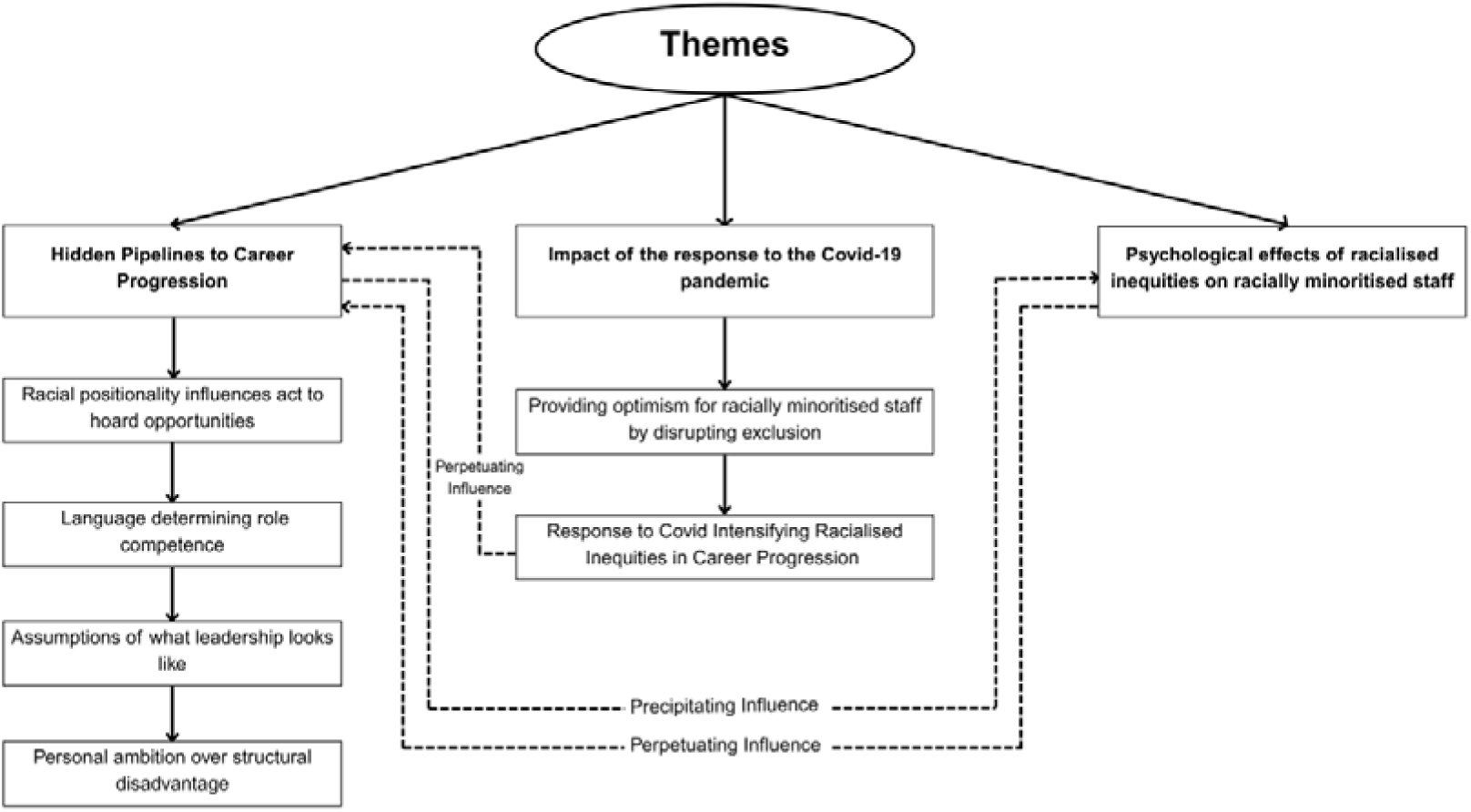
**Diagrammatic illustration of themes and subthemes.**

### 4.7 Ethical Considerations

Ethical approval was granted by the King’s College London Research Ethics Committee for Psychiatry, Nursing and Midwifery (HR-17/18-4629; RESCM-19/20-4629; RESCM-20/21-4629) and NHS Health Research Authority (18/HRA/0368).

### 4.8 Rigor and reflexivity

Data collection, data analysis and drafting of this paper were conducted by a research team with a diverse racial and ethnic composition. Most of the authors had extensive experience and knowledge in conducting qualitative research on inequities and inequalities in health services. As joint first authors, NMS and DW identify as Black early career researchers with previous experience collecting and analysing qualitative data. NMS has a background in inequities in health systems policy and DW has completed clinical placements as an assistant psychologist and is a postgraduate doctoral student in Clinical Psychology. Also of note, PM was a trained peer researcher with a nursing background who commented on sampling decisions, topic guides and interpretation of findings in regular TIDES project team meetings and contributed to paper drafts.

Rigorous discussions and meetings were held among the co-authors to review and refine the identified themes, ensuring interpretations of participants’ experiences were accurately represented in the data. Detailed research steps were documented during the study, including data collection, coding decisions, and analytical processes; the process and interpretation of findings were discussed in meetings with the national TIDES advisory and stakeholder opinion groups comprised of nurses, senior managers and other healthcare professionals.

## 5 Findings

### 5.1 Characteristics of participants

Data from 51 healthcare professionals across various specialities, roles and pay bands were included in the analysis (Table 1). Most participants identified as female (69%) and from the White (British and Other) group (41%). To preserve anonymity, we present race and ethnicity in our quotes as ‘Asian ethnicity’ (including Indian, Pakistani, Bangladeshi, Chinese and Other Asian groups), ‘Black ethnicity’ (including Black African, Black Caribbean and Other Black Groups), ‘White ethnicity’ (British, and Other White groups) and ‘Mixed/Other ethnicity’. Throughout this paper, participants from ‘White British’ and ‘Other White’ groups are referred to as ‘White staff’, while participants from ‘Asian or Asian British’, ‘Black or Black British’ and ‘Mixed/Other’ groups are referred to as ‘racially minoritised staff’. Gender and age are not presented alongside quotes to maintain anonymity.

**Table 1:** Sample characteristics.

|  | (n) | % |
| --- | --- | --- |
| **Gender** |  |  |
| Female | 35 | 68.6 |
| Male | 16 | 31.4 |
| **Race and ethnicity** |  |  |
| Asian or Asian British (Bangladeshi/<br>Chinese/ Indian/ Pakistani/Other Asian) | 13 | 25.5 |
| Black or Black British<br>(African/Caribbean/Other Black) | 14 | 27.5 |
| White (British/Other White) | 21 | 41.2 |
| Any other group (Mixed/Other) | 3 | 5.9 |
| **Role** |  |  |
| Student Nurse, Nurse, Nursing Associate<br>or Healthcare Assistant | 27 | 52.9 |
| Senior Staff or Senior Management | 24 | 47.1 |

### 5.2 Themes and subthemes

Three main themes and six subthemes were identified that highlighted how racial inequalities in career progression are maintained within the NHS (Figure 1). First, ‘*hidden pipelines to career progression*’ highlights how racial positioning influences unequal career trajectories shaped by informal networks, as well as racialised language, organisational norms and perceptions of competence. Second, ‘*impact of the response to the Covid-19 pandemic*’ shows how the pandemic disrupted and reinforced racialised career barriers depending on role and seniority. The third, ‘*psychological effects of racialised inequities on racially minoritised staff*’ captures the mental and emotional burden of navigating these inequities.

### 5.3 Hidden Pipelines to Career Progression

Participants’ accounts indicated that the route to career progression within the NHS was not transparent. They described a hidden pipeline that expedited career progression for White staff but excluded, or made more complicated, entry for racially minoritised staff. For participants, this pipeline was actively facilitated through networks that advantaged White staff since senior leaders – who often acted as gatekeepers to promotion and progression – were more commonly White themselves. Further, individual relationships between people within this network added value to career progression (e.g., offered opportunities to upskill). Here, Whiteness characterised the networks and led to in-group bias, based on shared personal attributes (Tajfel et al., 1971). Perspectives about this career pipeline varied, with White staff perceiving exclusion in terms of ambition rather than a racialised process.

#### 5.3.1 Racial positionality influences act to hoard opportunities

When asked about under-representation of racially minoritised staff in senior roles, racially minoritised staff across pay bands described the existence of informal networks (e.g., social circles) that provide access to key advice and opportunities. These networks were depicted as influential for career progression and consisted primarily of White staff.

> “The classic signs of snowy white peaks. You know, personal favours, it’s sad to say those things, but they are, they’re real, and they’re not very far from here.” Senior Leaders and Managers sample; Asian ethnicity

When describing these informal networks, racially minoritised staff often implied that exclusion was based on race. These networks were perceived as primarily serving White staff by providing greater access to development opportunities and pathways to higher-banded roles.

> “In the NHS is not what you know, it’s who you know…if your network is mainly of White counterparts then you progress and give your promotions to White counterparts which is why they are seen in upper management and senior management. That’s probably the reason I would attribute to it.” Post-Covid Sample; Black ethnicity

The accounts given by racially minoritised staff highlighted that team managers in these networks actively controlled access to progression opportunities. Managers positioned staff for promotion, such as offering new roles or opportunities to ‘act up’. This often resulted in White staff being invited and given access to opportunities for continued training and experience, or to progress into a new role, despite not being more experienced or qualified than their racially minoritised colleagues.

> “A White man, for example, that’s same grade as me hasn’t been at this grade for as long as I have, but will be invited to sit at the highest level of, the [NHS] Trust in terms of psychology and psychotherapy, leadership, and comment on things, and take on projects. And I don’t have that same exposure, or that same opportunity. So, there are those things’cause that will help him then to get to the next step up into an 8d job that he could go for, you know, because he’s had that experience and he’s got those networks. But for me I’m held back from some of those opportunities.” Senior Leaders and Managers sample; Black ethnicity

The inclusion of White staff and exclusion of racially minoritised staff from these informal networks suggested a form of opportunity hoarding, defined by Tilly (1999) as a method of exclusion. Methods of exclusion from the informal networks were highlighted by some racially minoritised staff as being based on cultural attributes (e.g., language, norms and values). Such attributes were perceived as negative, or in the case of senior racially minoritised staff, as not fitting in with organisational norms and a culture of Whiteness. The mechanisms underpinning exclusion from the informal networks reflect the idea of Whiteness as a credential and are illustrated by the subthemes ‘*Language determining role competence’* and ‘*Assumptions on what leadership looks like*’.

#### 5.3.2 Language determining role competence

When discussing hurdles for career progression and reasons behind the lack of representation, some racially minoritised staff discussed the importance of English language dialect and proficiency, as this often influenced perceptions around competence. Expectations regarding how staff speak often reflected a norm of Whiteness, how staff are ‘expected’ to sound at a senior level. When asked about ethnic inequities in career progression, one participant highlighted issues around language skills, and centred tackling this as a solution.

> “…here based at [NHS Trust] we get lots of our junior managers coming from the local area, but actually having grown up within a specific part of London speaking like each other, that doesn’t win you very many,-or it doesn’t allow you to be acknowledged [for career progression] sometimes if you’re not able to speak in the way that you’re expected to speak. And it alienates you from kind of people in those higher-ranking posts. So, it is kind of interesting how you learn to talk in a different way, don’t you? When you’re in a certain kind of meeting versus a different kind of meeting. So, I think the language skills is a really important thing that we just don’t focus enough on. You know, correct grammar, correct approach to writing an email you know, all the kinds of things that actually change the way that people view you.” Senior Leaders and Managers sample; Asian ethnicity

English language dialect and proficiency were viewed as a social asset that implied stronger leadership qualities for those racially minoritised staff who were able to “learn to talk in a different way”. This appeared to shape the boundaries of informal networks and contributed to the exclusion of racially minoritised staff from career progression.

#### 5.3.3 Assumptions of what leadership looks like

Racially minoritised staff recounted experiencing hostility and exclusion from colleagues, particularly when entering positions of leadership or high responsibility. These experiences were described by racially minoritised staff as being a result of their race, and how it did not align with the organisation’s norms around leadership.

> “When we occupy positions of leadership within healthcare services, the message that we get from our colleagues is that we shouldn’t occupy these positions…So I’ve worked as a [leadership role] in a number of services, and I’ve had different experiences of how services might receive me as a BME [Black and minority ethnic] leader. When I came to this [NHS] Trust, one of my first felt sense was that I was highly different, and that my difference was a problem. I wasn’t able to kinda name it…But it felt-my difference-the fact that I looked foreign was a problem for individuals…” Senior Leaders and Managers sample; Asian ethnicity

The nature of hostility experienced by racially minoritised staff suggested that microaggressions, as described by Williams (2019) as “deniable acts of racism that reinforce stereotypes and inequitable social norms” (p.4), acted to enforce hierarchical boundaries through stigmatisation. Racially minoritised staff were implicitly told that they were not welcome in leadership spaces at work.

#### 5.3.4 Personal ambition over structural disadvantage

Experiences of exclusion described by racially minoritised staff when discussing racialised inequalities in career progression were not mentioned by many White staff. Some White staff attributed lack of progression to low confidence or aspiration among racially minoritised staff.

> “I don’t think people are leaving necessarily, I just don’t think that they chose to go up the ladder to more experienced posts…we’ve carried on doing quite a lot of recruitment at senior levels. Interestingly enough, we went out for an advert recently, for three posts, we had 21 applications, we shortlisted seven, not one person was BME. So it’s not that we, you know what I mean, but they’re just not applying.” Senior Leaders and Managers sample; White ethnicity

When asked about the racialised inequity in career progression, several White staff dismissed the impacts of discrimination and the advantage it gives to White staff groups. Instead, they emphasised a framework where progression depended solely on personal ambition.

> “I think it’s down to the individuals themselves, purely because if you want to achieve something, you have to put something in it. You have to show the willingness of wanting to learn, wanting to do something new, wanting to progress […] I look at everybody as equal,’cause we’re given equal opportunities to do one thing or another, to learn, to progress. It’s just that you wanting and you’re willing to do it…We have such a vast diversity of-mix of people working so it’s impossible to even try-even think of discriminating one or another.” Post-Covid sample; White ethnicity

In contrast, certain White staff mentioned the presence of informal networks and acknowledged that these facilitated opportunities for career progression. However, their perspectives differed from racially minoritised staff, since entry into these networks was not perceived as a systemic racialised process but based on individual likeability or personal characteristics. The lack of focus on Whiteness was evident when discussing the role of networking in career progression.

> “In many places…not just the current workplace-there are always jobs being created for particular person. Usually, they are a person who are in a good relationship with a very senior [person]…whether that’s favouritism for the individuals or because of their personality, or some other reasons such as race, sexuality, I think they play a part in it as well. But recruitment isn’t as open and transparent as it should be. I’ve certainly been on the receiving end of that and witnessed it numerous times so…yeah…I think it’s the person that best gets on with whoever is doing the recruitment…It’s still a bit of ‘jobs for the boys’ or ‘the girls’, so to speak. It’s better, but it’s still…there’s ways around, there’s ways around the recruitment procedure to manipulate that.” Senior Leaders and Managers sample; White ethnicity

This reluctance to address Whiteness appeared to minimise the possible institutional privileges White staff may have for career progression.

### 5.4 Impact of the Response to the Covid-19 Pandemic

The Covid-19 pandemic was a macro-level force that impacted informal networking in the NHS since it altered ways of working (e.g., through redeployments and remote working). This in turn affected how career progression opportunities were accessed and distributed. These changes were described by participants with both optimism and negativity but were not patterned by ethnicity.

#### 5.4.1 Providing optimism for racially minoritised staff by disrupting exclusion

Those who optimistically described the impact that Covid-related changes in working had on their career progression were mostly non-senior staff and newly qualified nurses. These participants highlighted how redeployment increased their exposure to different environments and, more importantly, different skillsets.

> “With the pandemic a new service was created in our-in our Trust which is provided, I would say better opportunities for career progression for nurses. So they created a sort of emergency assessment unit. Where-to reduce the flow of patients to A&E [Accident and Emergency]. It was sort of our own little psychiatric liaison, and so maybe it’s given me more opportunities within my Trust and so I’ve done some overtime shifts that so maybe it could help enhance my career progression.” Post-Covid sample; Black ethnicity

The new access to opportunity resulting from redeployment seemed to thwart the status quo by providing racially minoritised staff the opportunity to develop skillsets and advance their careers. With this, some non-senior nursing staff were able to bypass the gatekeeping of opportunities to strengthen their competencies.

#### 5.4.2 Response to Covid Intensifying Racialised Inequities in Career Progression

Contrastingly, disruption to ways of working experienced by some non-senior staff was not mirrored by more senior staff. Their redeployments gave no new opportunity to develop skills and were seen as disruptive to career progression.

> “In terms of some of the development programmes, we are seeing Covid impacting a bit where…people that are on these programmes are clinical again.. or be redeployed. So that might be slowing down the sort of career development opportunity that might be there if it was just sort of normal. You know, pre Covid.” Senior Leaders and Managers sample; Asian ethnicity

Other changes to work style, such as remote working, were viewed by racially minoritised staff as adversely impacting career progression due to gaps in CVs and strained relationships with managers. This was particularly true for minoritised staff who were clinically extremely vulnerable to COVID and were advised to shield (stay home and minimise face to face contact with others).

> “I was discouraged from working from home because I wouldn’t be able to support [my colleagues] and it may look bad on my career in the future…I was very forthright and said, “Well, I’ve got a shielding letter and she hasn’t, so I will be working from home,’cause here’s the government’s letter.” But that really did worry me because I’m gonna have to ask my manager for a reference and I have no clue what [they’re] going to put on that reference. Or then do I take an 8 month gap out of my CV so that I can negate [them] giving me a bad reference? I’ve worked in healthcare since I was 18, and so it’s worrying to think, what [are they] gonna write on this? Because that will stop me from moving forward.” Senior Leaders and Managers sample; Black ethnicity

Similarly, lockdown and social distancing measures meant that some could not physically access opportunities.

> “Obviously at the moment it’s all just been about reviewing patients or supporting other teams. So, in terms of opportunity, there’s obviously been less.’cause you’re physically not obviously going out doing teaching or you know, having meetings. Or you know, writing publications, that kind of thing. I’d say that’s how it’s impacted. There’s kind of been less developmental opportunities, but I wouldn’t say there hasn’t been any. Um, it’s just not been the same.” Senior Leaders and Managers sample; Asian ethnicity

These disruptions to work patterns of senior racially minoritised staff appeared to compound the gatekeeping perpetuated by managers (Figure 1), since access to development opportunities became even more limited.

### 5.5 Psychological effects of racialised inequities on racially minoritised staff

Despite ‘working hard’ and applying for positions, racially minoritised staff expressed that they could not progress in their careers. This lack of progression and exclusion from the informal networking negatively impacted their emotional wellbeing and contributed to racial battle fatigue, defined as psychosocial stress responses to ongoing exposure to racism and discrimination as a member of an oppressed group (Smith, 2008).

> “I don’t have the energy to do any extra things to prove to people that I’m equally as good as them, or I’m equally as good as my peers who are White…Yeah…and I feel defeated by this. And I don’t have the energy to fight it anymore” Senior Leaders and Managers sample; Asian ethnicity
>
> “I did a Masters and then I went into the NHS and even now I still think I don’t get as many opportunities as some of my other colleagues. And that’s challenging’cause I have more experience and more *academic* skills, as in compared to some of my colleagues and that has knocked my confidence because I do often think about leaving the NHS. Especially this last year has really challenged me and I don’t know if that’s because I’m working in a hospital I’m the only ethnic minority at band 8b and it’s the way it’s been, how some people have communicated with me has been such a negative experience.” Senior Leaders and Managers sample; Black ethnicity

The fatigue appeared to be a product of the accumulated exposure to discrimination at each stage of the career ladder. Repeated exclusion from career progression opportunities led racially minoritised staff to feel compelled to leave their role due to ongoing sexposure to these stressors.

> “It’s a lot harder for people from these [racially minoritised] backgrounds to actually be able to get to those [senior roles], because they’re not exposed to the opportunities that are available and you know, there’s a lot of they have to work 10 times harder to get to those roles. And I think there’s always an issue once they get there, there’s a lot of frustrations and they can’t really stay on top for a really long time. From the conversations I’ve had with different people […], that’s always the biggest, not having those opportunities available, and when they do come, there’s a lot of difficulty and hurdles for you to actually maintain that position and then somehow somewhere down the line you just have to leave, you know, get away from that position or the stress is just overwhelming that you can’t really stay in that position for too long.” Post-Covid sample; Black ethnicity

For racially minoritised staff, repeated discrimination also appeared to lead to anticipated discrimination.

> “And I suppose that becomes self-perpetuating, because if you have heard about these experiences of your people who look like you, then you’re then the expectation is that it’s going to happen to you too, so you think I don’t have the energy to put myself through that, so you’re likely to talk yourself out of having to be exposed to that, particularly if you’re carrying some form of that anyway.” Senior Leaders and Managers sample: Black ethnicity

With this, racially minoritised staff conveyed that they felt like giving up, or saw their counterparts give up. This process had an implicit underlying message – that racially minoritised staff do not belong in senior positions.

> “I’ve got experience in my service where my White colleagues were allowed to access CPD [continuing professional development] to progress. And when I asked for something similar, it was declined. And that consistently happens to BME staff-where their White colleagues will be given kinda opportunities to develop, to learn-but not to BME staff…And there’s no genuine reason given for that, apart from, ‘Well, it doesn’t fit with the service needs at the moment’….And often, it becomes like for BME staff-‘Which battle do I want to fight?’, I am not kidding you, it’s exhausting….It is mentally, psychologically, physically exhausting and staff give up. A lot of BME staff feel completely burnt out!…And then they stop feeling motivated to learn, or to progress-‘If my senior managers don’t see me as a leader, if my colleagues don’t see me as a leader, I’m less likely to apply for that post that comes up that’s a senior post’. Senior Leaders and Managers sample; Asian ethnicity

The internalisation of this messaging often damaged confidence and ability of racially minoritised staff to progress (Figure 1). This internalisation and anticipated discrimination may inhibit staff from pursuing higher banded roles, in contrast to White staff perspectives of inequity as a ‘lack of ambition’.

## 6 Discussion

Our findings highlight that access to opportunities for career progression in the NHS is neither transparent, fully democratised nor based on merit. Racial inequities in career progression are maintained through informal networks that hoard opportunities. These networks benefit White staff by giving them greater proximity to managers who gatekeep access to opportunities. Managers may also exclude racially minoritised staff by stigmatising their cultural attributes. Moreover, the response to the Covid-19 pandemic has impacted the process of exclusion resulting from informal career-advancing networks. While non-senior racially minoritised staff perceived increased access to opportunities, senior racially minoritised staff experienced a heightened sense of exclusion. Our findings demonstrate how racialised inequity in career progression is perpetuated, at least partly, by exclusion of racially minoritised staff from informal networks. We also illustrate how lack of career progression adversely affects psychological state of racially minority staff, expanding on the findings of Rhead et al. (2020).

The frequent rejections and exclusions were depicted by some racially minoritised staff as leading to a feedback loop whereby fatigue led to a desire to give up on career progression, thus rendering the inequity in career progression a self-fulfilling prophecy (Figure 1). Such resignation was incorrectly labelled by White staff as a’lack of ambition’. This labelling shifted blame onto racially minoritised staff groups rather than the inequality-generating actions of advantaged White staff groups, an example of a health inequality diversion (Link and García, 2021). This is important to address since the racialised inequity in career progression, in addition to impacting staff retention, limits knowledge and expertise in service delivery. These racialised inequities affect staff morale and wellbeing, adversely affect patient care, and are reputationally damaging for the NHS (Rhead et al., 2021; Todorova et al., 2010; Woodhead et al., 2022).

### 6.1 Strengths and Limitations

Our study supports theory around racialised organisations and diversions addressing racial discrimination. A strength is that it takes perspectives from staff nationally during Covid-19 pandemic restrictions. This gives the study the real-time perspectives of the impact the response to the pandemic had on exclusion of racially minoritised staff across workplace networks. However, this could also be a limitation, as there was little time for staff interviewed in this study to process their experiences as NHS staff during the height of the pandemic. It is possible that staff could not completely see how Covid-19 impacted exclusionary processes, and it is unclear whether staff would have similar views and experiences post-pandemic. Therefore, a follow-up study or extending our longitudinal approach would allow us to examine changes and impact of the pandemic on racial inequalities in career progression and retention.

### 6.2 Recommendations for Further Research

#### 6.2.1 Whiteness as a credential in the NHS

Participants’ accounts described an informal, racially biased pipeline for career progression that was reliant on the discretion of, and proximity to, managers and other beneficial relationships through informal networks. Evidence from Dhaliwal et al. (2017) and Isaac (2020) has highlighted how differences in social or cultural capital can inhibit progression in the NHS for racially minoritised staff. In this paper, we defined social capital as the value of social network relationships, which provide access to benefits and solutions to problems through reciprocity (Bourdieu, 1986; Sander and Lowney, 2006) and cultural capital as acquired social assets which enable progress in a stratified society (Bourdieu, 1986; Barker, 2004). Our findings indicate that social capital has a role in NHS career progression through informal networks providing access to benefits and resources such as promotion, training and career development. Moreover, Isaac (2020) describes how the acquisition of ‘British cultural capital’ by Black mental health nurses allowed them to shape their careers by mitigating discriminatory bias. Our findings build on this point by suggesting that cultural attributes of Whiteness, such as accent and language proficiency, are actively centred and valued by managers in the informal networking groups, giving those with the cultural capital of Whiteness the currency to enter. Evidence from Woodhead et al. (2022) helps illuminate why the cultural capital of Whiteness acts as such currency, since it was suggested that the organisational layers of culture (the ways things are done, shared ways of thinking and deeper shared assumptions) in the NHS are inherently racialised. In support of the third tenet of Ray’s theory of Racialised Organisations (2019), our findings suggest that Whiteness, in the form of cultural capital, acts as a credential within the NHS. This grants entry into informal networking groups, facilitating relationships necessary for progression.

#### 6.2.2 Denying and displacing structural advantage

Lack of attention to the role of Whiteness in career progression, particularly from White staff, can be applied to Link and García’s (2021) conceptualisation of inequalities since it posits that the career advantage White staff have is partially maintained by lack of its acknowledgement. Our findings propose that maintaining this advantage may perpetuate deficit narratives, which lay the blame for the racialised inequity in career progression on those it affects most. Applying these findings to Link and García’s (2021) conceptualisation suggests that diverting attention from White staff to racially minoritised staff stops scrutiny on Whiteness and the advantage it gives to White staff in career progression. The result is the continuation of opportunity hoarding for White staff within the informal networks, as interventions are focussed on individuals and directed at racially minoritised staff. Through this interpretation, our findings expand on Woodhead et al. (2022) by showing how a White majority organisation, in this case the NHS, can maintain advantage for White staff.

#### 6.2.3 Impact of the Covid-19 response

The impact of the response to the Covid-19 pandemic provided optimism for non-senior staff members’ perceptions about career progression. These staff felt that the exclusion they experienced was overcome by new access to opportunities that could further their career. However, senior staff saw the response to Covid-19 as an event that maintained the exclusion of racially minoritised staff. This raises the question: will non-senior racially minoritised staff see the more equitable progression that they themselves have requested? The pandemic threatened long-established ways of working underlying inequalities, evidenced by micro-aggressions (Sue et al., 2007) experienced by senior racially minoritised staff when entering senior spaces. These experiences highlighted hostility racially minoritised staff face for entering a space in which they are typically underrepresented. Further research needs to examine how changes in work have impacted the careers of racially minoritised staff at varying levels of seniority, including clinical and non-clinical staff. Longitudinal quantitative and qualitative studies are needed to ascertain whether there were any racialised changes in career trajectory from the first UK-wide Covid-19 lockdown, if any changes were sustained, and the mechanisms underlying these patterns.

### 6.3 Implications for Policy and Practice

There have been efforts to de-bias the recruitment process, such as blind recruitment. However, data from the Workforce Race Equality Standard (WRES) 2024 report suggests such efforts have not yet equalised progression. Despite some small regional improvements in recruitment and promotion, White staff are still more likely to be appointed to roles via shortlisting and to be the appointing managers (NHS England, 2023). Accounts from participants further suggest that NHS Trusts’ efforts to address inequity in career progression have not fully tackled problems with recruitment and promotion since (1) informal networks benefit White staff at the expense of racially minoritised staff; and (2) exclusion from these networks can lead to anticipated discrimination that inhibits racially minoritised staff from pursuing higher banded roles. Subsequently, actions towards addressing racialised inequities in career progression need to be cognisant of the advantage White staff hold and pervasive deficit narratives. Transparently acknowledging and owning these two points within organisations provides fertile ground for existing and new approaches to eliminating racialised inequities in career progression in the NHS.

Changing from a deficit narrative to one of structural advantage can be enacted through staff training. Staff could be made more aware of what Whiteness is, how it manifests within the NHS as a bias, and how this bias influences the exclusion of racially minoritised staff from informal networking groups because of their racial identity and cultural attributes. Learning from the ‘Improving Race Equality in Health and Social Care’ report for the Wales Centre for Public Policy (Hatch et al., 2021), and the ‘Developing Anti-Racist Practice To Support Black And Other Racial Minority Nurses And Midwives Within The NHS’ review for the Chief Nursing Officer (Jieman et al., 2022), such staff training would integrate an anti-racism lens that challenges Whiteness as the institutional norm and associated privileges within the NHS. A core aim would be to increase understanding of how racism impacts inequities in career progression and access to training and opportunities. To do so, Hatch et al. (2021) suggest that this training needs to be action-oriented and an ongoing feature of staff development, with built-in accountability processes. Delivery of this training needs to be spread across all NHS staff and not just focussed on racially minoritised staff. This is to help all staff increase their understanding and confidence when tackling racial inequities within the workforce, and to also help understanding of how tackling racial inequities benefits everyone (Robertson et al., 2021).

It is crucial that this training is mandatory throughout the workplace hierarchy; although it was identified by staff that managers actively controlled progression, exclusion was experienced by racially minoritised staff in both the senior and non-senior samples. Inclusion as a practice needs to be integrated across the career trajectory from the student and early career stage to the director level, and senior leadership would have a critical role in shaping this (Jieman et al., 2022; Ross et al., 2020). Insights from the King’s Fund and The NHS Race and Health Observatory show that when senior leaders lead by example, they help to convey the importance of tackling racial inequities within the workforce (Ross et al., 2020). Leadership practice, leadership development and team-working interventions need to acknowledge the influence of senior leaders to help cultivate and maintain universally inclusive practices (Ross et al., 2020; Jieman et al., 2022). The desired outcome of these proposals is to further embed inclusivity within the NHS organisational culture and the career progression pipeline. To better understand and address the psychological impact of racialised hierarchies within the NHS, it is recommended that psychological support – either through one on one or group interventions – be made available to all racially minoritised staff.

To complement staff training proposals, there should be a commitment to further research on organisational culture within healthcare systems, with an increased focus on how groups are advantaged. This includes how they benefit, how they maintain such benefit and who they disadvantage. Future research should consider potential differences in the experiences of British-born and internationally recruited NHS staff who may have distinct forms of social and cultural capital. Qualitative methodology should be used to enhance existing quantitative data from staff surveys to better understand mechanisms behind the inequities. There should also be formal evaluations of how existing interventions, such as positive or proportionate action interventions which allow an employer to proportionately reduce the disadvantage, meet needs or increase participation of persons from a protected group (Equality Act, 2010) to accelerate the reduction of racial inequities in career progression. We need to question how those interventions have been performing historically, and whether they operate in a deficit narrative framework. Such appraisals should be undertaken on a Trust-by-Trust basis with regional comparisons, since geographical and social context will be important.

## 7 Conclusion

NHS staff perspectives on racialised inequities in career progression highlight the power of informal networks for giving staff access to managers who control opportunities for progression. These networks appear to operate within a culture of Whiteness, excluding racially minoritised staff and advantaging White colleagues. Repeated exclusion and stalled progression have negatively impacted the psychological well-being of many racially minoritised staff, leading to fatigue, pessimism and reduced staff retention – ongoing issues in the NHS. While some staff found new opportunities during the Covid-19 response, others, particularly senior racially minoritised staff, felt redeployments and remote working hindered their development. Notably, many White staff, both senior and junior, appeared unaware or dismissive of how Whiteness shaped these inequities. To address this, the NHS must recognise and actively challenge how Whiteness as an organisational culture maintains racialised inequity in career progression. Without this, solutions risk defaulting to deficit-based approaches that only focus on the perceived faults of racially minoritised staff, leaving structures, processes and norms that perpetuate racialised inequities in career progression in place.

## Data Availability

Data are available from the Kings College London Figshare repository: TIDES Phase 2 Interview Study, doi:10.18742/22101953. Access is subject to the Kings Data Access Agreement.

https://doi.org/10.18742/22101953

